# Nonlinear relationship between platelet count and 30-day in-hospital mortality in ICU stroke patients: a multicenter retrospective cohort study

**DOI:** 10.1101/2024.01.14.24301300

**Authors:** Lan-xiang Wang, Ren-li Liu, Pan Zhou, Hao-fei Hu, Zhe Deng

## Abstract

**Background:** Evidence of the relationship between platelet count and 30-day in-hospital mortality in ICU stroke patients is still scarce. Therefore, the purpose of this study was to explore the relationship between platelet count and 30-day in-hospital mortality among ICU stroke patients.

**Methods:** We conducted a multicenter retrospective cohort study using data from 8029 ICU stroke patients in the US eICU-CRD v2.0 database from 2014 to 2015. Utilizing binary logistic regression, smooth curve fitting, and subgroup analyses, we examined the link between platelet count and 30-day in-hospital mortality.

**Results:** The 30-day in-hospital mortality prevalence was 14.02%, and the mean platelet count of 223×10^9^/L. Adjusting for covariates, our findings revealed an inverse association between platelet count and 30-day in-hospital mortality (OR = 0.975, 95% CI: 0.966, 0.984). Subgroup analyses supported the robustness of these results. Moreover, a nonlinear relationship was observed between platelet count and 30-day in-hospital mortality, with the inflection point at 163×10^9^/L. On the left side of the inflection point, the effect size (OR) was 0.92 (0.89, 0.95), while on the right side, the relationship was not statistically significant.

**Conclusion:** This study establishes an independent negative association between platelet count and 30-day in-hospital mortality in ICU stroke patients. Furthermore, a nonlinear relationship with a saturation effect was identified, suggesting that maintaining the platelet count around 163×10^9^/L can reduce 30-day in-hospital mortality in these patients.

## 1. Introduction

Stroke, the second leading global cause of death and third in disability(1, 2), recorded 12.2 million incidents, 101 million prevalent cases, and 6.55 million deaths in 2019(2). In the U.S., nearly 800,000 patients are affected annually, with a rising proportion of young stroke patients(3, 4). Despite recent progress in treatment, the global burden of stroke is rapidly increasing, posing challenges many countries can’t meet(1). In summary, stroke significantly impacts social economy and health(5), necessitating early identification of risk factors for poor patient outcomes.

While studies identify factors like age, obesity, diabetes, hypertension, heart disease, and stroke etiology affecting stroke outcomes(2, 6–8), intervening in these factors during a stroke is challenging. Thus, some studies focus on the relationship between laboratory biomarkers and stroke outcomes(9, 10), as these factors are more manageable.

Platelets, produced by 2-4 μm megakaryocytes, circulate for 7-10 days, primarily responsible for hemostasis and coagulation(11). Recent research links platelets not just to thrombosis but also other physiological and pathological processes. Studies associate platelet count and mean platelet volume with tumor metastasis(12, 13). Additionally, within the normal range, elevated platelet count correlates with increased arterial stiffness in young and middle-aged individuals(14). A retrospective analysis of 2056 intensive care units (ICU) patients associate thrombocytopenia with acute respiratory distress syndrome and death(15). In a prospective study of 84 critically ill patients, elevated platelet volume after ICU admission independently associates with higher in-hospital mortality(16). However, the association between platelet count and stroke outcomes lacks epidemiological evidence. Considering the unique physiological basis of critically ill patients, we speculate a decreased baseline platelet count before treatment may relate to poor stroke prognosis. High-quality multicenter data clarifying this relationship can offer reliable evidence for future studies.

Thus, we analyzed the eICU-CRD v2.0 to explore the impact of baseline platelet count on 30-day in-hospital mortality in ICU stroke patients.

## 2. Methods

### 2.1. Study Design

This study is a multicenter retrospective cohort, focusing on platelet count as the independent variable and 30-day in-hospital death in stroke patients as the dependent variable (dichotomized into death or survival).

### 2.2. Data Source

The eICU Collaborative Research Database, a freely available multi-center database for critical care research. Pollard TJ, Johnson AEW, Raffa JD, Celi LA, Mark RG and Badawi O. Scientific Data (2018). DOI: http://dx.doi.org/10.1038/sdata.2018.178. Available from: https://www.nature.com/articles/sdata2018178. This database comprises high-quality data from 200,859 patients admitted to ICU in 208 U.S. hospitals during 2014 and 2015. It includes various clinical information such as vital signs, nursing plans, disease severity, diagnosis, and treatment details(17). The data is accessible to the public after registration, with the collection process meeting the standards of the MIT Ethics Committee (No: 0403000206) and aligning with the 1964 Declaration of Helsinki.

### 2.3. Study Population

This study included 8209 eligible individuals, determined by the following exclusion criteria: (1) non-stroke patients: n=189,752 excluded; (2) age<18 years: n=3 excluded; (3) ICU stay time <24 hours or >30 days: n=2436 excluded; (4) missing in-hospital mortality data: n=109 excluded; (5) missing platelet count: n=246 excluded; (6) extreme values (three standard deviations above or below the mean) of platelet count: n=104 excluded. After applying these criteria, 8209 participants remained for the analysis. The participant selection process is illustrated in Figure 1.

**Fig 1.**
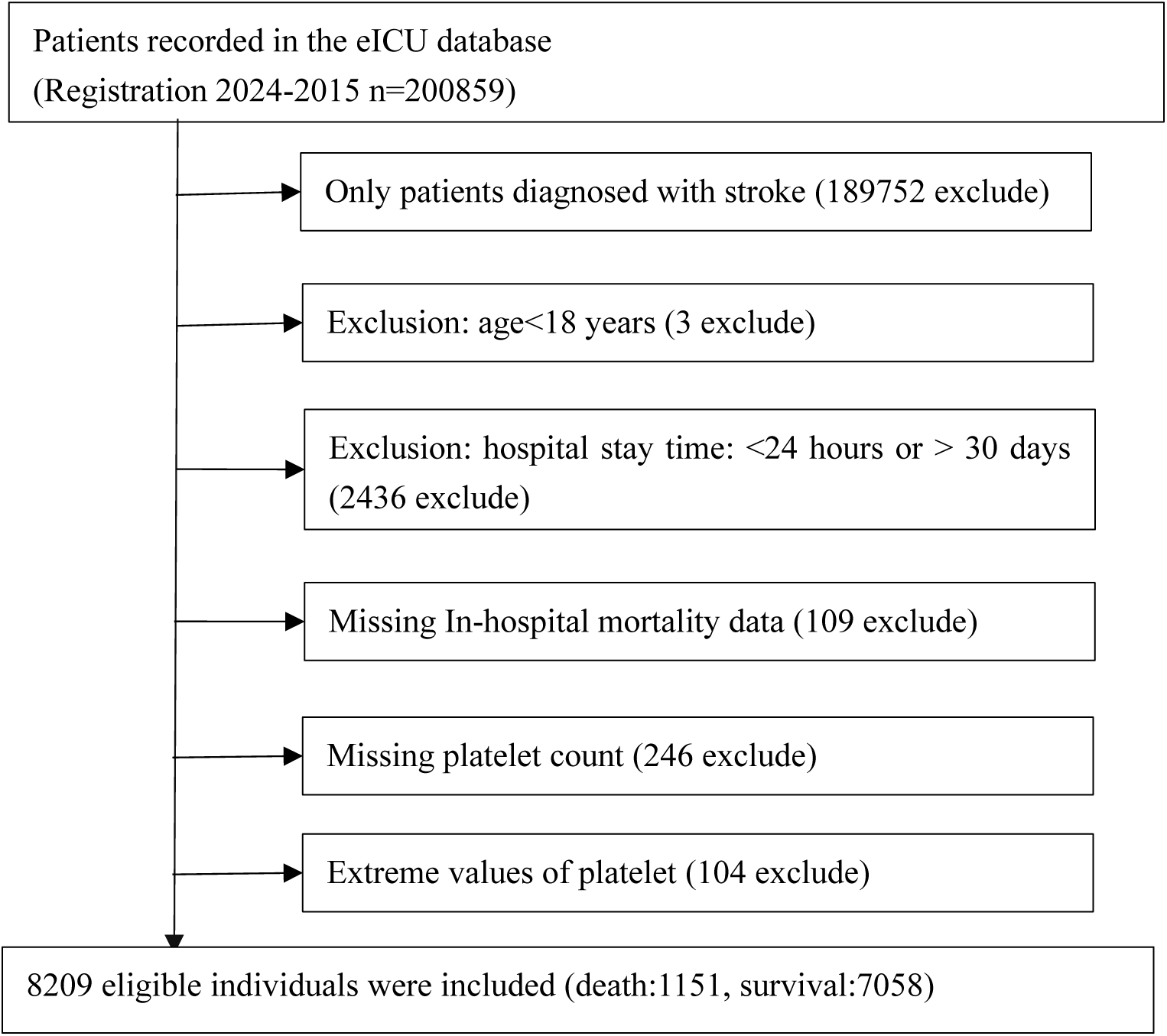
Flowchart of study participants

### 2.4. Variables

The platelet count was initially recorded as a continuous variable. Subsequently, it was categorized based on quartiles: Q1 (2.0-172.0×10^9^/L), Q2 (173.0-215.0×10^9^/L), Q3 (216.0-264.0×10^9^/L), and Q4 (265.0-487.0×10^9^/L).

### 2.5. Covariates

The covariates were selected based on clinical experience and previous studies(18–21). The included variables were: (1) categorical variables: sex, ethnicity, acute respiratory failure (ARF), atrial fibrillation (AF), acute coronary syndrome (ACS), congestive heart failure (CHF), chronic kidney disease (CKD), chronic obstructive pulmonary disease (COPD), diabetes mellitus, gastrointestinal bleeding (GB), hypertension, sepsis, use of anti-platelet medications, anticoagulants, glucocorticoids, carbapenems, cephalosporins, vancomycin, and mechanical ventilation; continuous variables: age, body mass index (BMI), hemoglobin concentration (Hb), serum creatinine (Scr), and Acute Physiology and Chronic Health Evaluation-IV score (APACHE-IV score). BMI was computed as weight (kg) divided by height (m) squared (kg/m^2^). Baseline parameters were collected within 24 hours of ICU admission.

### 2.6. Missing data processing

In the present study, the number of participants with missing data of sex, BMI, Scr and APACHE-Ⅳ score were 1(0.01%), 99(1.21%), 33 (0.4%), and 980 (11.94%), respectively. Multiple imputations handled missing covariate data(22), with the imputation model encompassing ethnicity, age, Hb, ARF, AF, ACS, CHF, CKD, COPD, diabetes mellitus, GB, hypertension, sepsis, use of anti-platelet medications, anticoagulants, glucocorticoids, carbapenems, cephalosporins, vancomycin, and mechanical ventilation. The missing data analysis assumption was based on the missing-at-random assumption (MAR) (23).

### 2.7. Statistical analysis

Individuals were categorized into four groups based on platelet count quartiles. Continuous variables were presented as mean (standard deviation) for normally distributed data and median (range) for non-normally distributed data. Categorical variables were represented as numbers (%). To assess differences between platelet count groups, we employed one-way analysis of variance for normal distribution, the χ2 method for categorical variables, or the Kruskal–Wallis H test for skewed distribution.

#### 2.7.1. To analyze the independent linear relationship of the platelet count and 30-day in-hospital mortality

After collinearity screening (Table S1: no covariates were excluded), three models were constructed using univariate and multivariate binary logistic regression, in accordance with the STROBE statement (24), to examine the relationship between platelet count and 30-day in-hospital mortality. The models were: (1) crude model (no covariates adjusted); (2) minimally-adjusted model (model I: adjusted for sex, age, and ethnicity); fully-adjusted model (model II: adjusted for sex, ethnicity, age, BMI, Hb, Scr, APACHE-IV score, ARF, AF, ACS, CHF, CKD, COPD, diabetes mellitus, GB, hypertension, sepsis, anti-platelet, anticoagulant, glucocorticoid, carbapenems, cephalosporins, vancomycin, and mechanical ventilation). Effect sizes with 95% confidence intervals (95% CI) were recorded. Adjustments for covariates were made when their addition to the model resulted in an odds ratio (OR) change of 10% or more (24). Subgroup analyses were conducted to assess result robustness. Platelet count was categorized by quartile, and P for trend was calculated to test continuous variable results and explore non-linearity. Additionally, potential unmeasured confounding between platelet count and 30-day in-hospital mortality was investigated using E-values.

#### 2.7.2. Subgroup analysis

Subgroup analyses utilized a stratified binary logistic regression model across various subgroups (including sex, ethnicity, ARF, AF, ACS, CHF, CKD, COPD, diabetes mellitus, GB, hypertension, sepsis, and mechanical ventilation). Besides the stratification factor, each stratum was adjusted for all factors (sex, ethnicity, age, BMI, Hb, Scr, APACHE-Ⅳ score, ARF, AF, ACS, CHF, CKD, COPD, diabetes mellitus, GB, hypertension, sepsis, anti-platelet, anticoagulant, glucocorticoid, carbapenems, cephalosporins, vancomycin, and mechanical ventilation). Finally, interaction tests were conducted using the likelihood ratio test of models with and without interaction terms (25, 26).

#### 2.7.3. To analyze the nonlinear relationship of the platelet and 30-day in-hospital mortality

Utilizing binary logistic regression models can sometimes be limited in addressing nonlinearity. To address this concern, we applied generalized additive models (GAM) and employed smooth curve fitting (penalized spline method) to further investigate the nonlinear relationship between the platelet count and 30-day in-hospital mortality. In the presence of nonlinearity, our approach involved initially determining the inflection point through a recursive algorithm. Subsequently, we established a two-piece binary logistic regression model on either side of the inflection point (27). The log-likelihood ratio test was then employed to identify the most appropriate model that describes the association between the platelet count and 30-day in-hospital mortality.

All results adhere to the STROBE statement (24). Analyses were conducted using statistical software packages R (R Foundation)2 and EmpowerStats3 (X&Y Solutions, Inc., Boston, MA). Statistically significant findings were determined at a threshold of *P* < 0.05 (two-sided).

## 3. RESULTS

### 3.1. Characteristics of Individuals

The individuals’ characteristics are summarized in Table 1. Analysis of 8209 patients based on platelet count quartiles (Q1-Q4) revealed notable trends in various demographic and clinical features. Significantly higher 30-day in-hospital mortality was observed in the lowest quartile (P<0.001). Male sex demonstrated a significant prevalence in lower quartiles compared to higher ones (P<0.001). Age exhibited a significant decrease across quartiles (P<0.001), suggesting a correlation between older age and lower platelet counts. Ethnicity, however, did not significantly differ between quartiles (P=0.177). Clinically, lower platelet quartiles were associated with lower BMI (*P*<0.001), lower Hb (P<0.001), higher serum creatinine (P<0.001), and higher APACHE-IV scores (P<0.001), indicating a potential connection between lower platelet counts and worse clinical status. Several comorbidities, including ARF (P<0.001), AF (P<0.001), CKD (P=0.049), and sepsis (P<0.001), exhibited significant differences between quartiles, suggesting that lower platelet counts may correlate with an increased risk of these complications. Notably, the usage of certain antibiotics like cephalosporins (P=0.005) and vancomycin (P=0.004), as well as the need for mechanical ventilation (P<0.001), were higher in lower platelet quartiles.

Figure 2 depicts the distribution of platelet count, presenting a normal distribution within the range of 2.0×10^9^/L to 487.0×10^9^/L, with a mean of 223.0×10^9^/L. In Figure 3, participants were categorized into two groups based on survival status. The platelet count distribution in the death group was notably lower, while the survival group exhibited a relatively higher platelet count.

**Fig.2.**
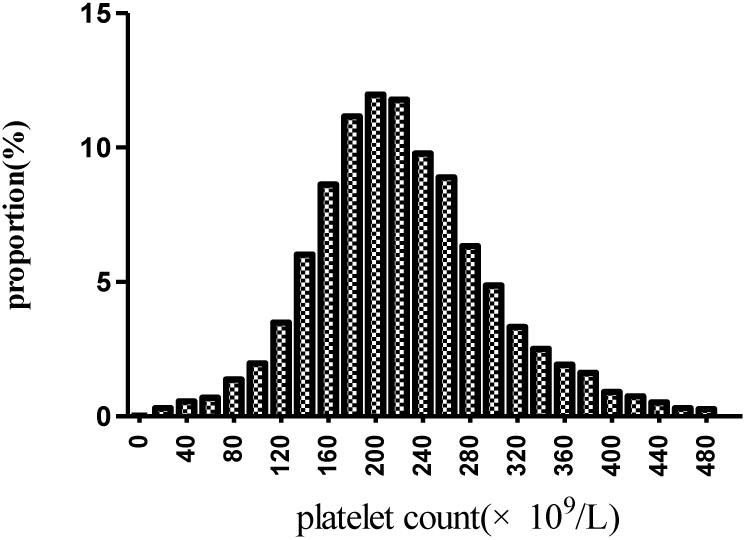
Distribution of platelet count. It displayed a nearly normal distribution ranging from 2×10^9^/L to 487×10^9^/L, with an mean of 223×10^9^/L.

**Fig. 3.**
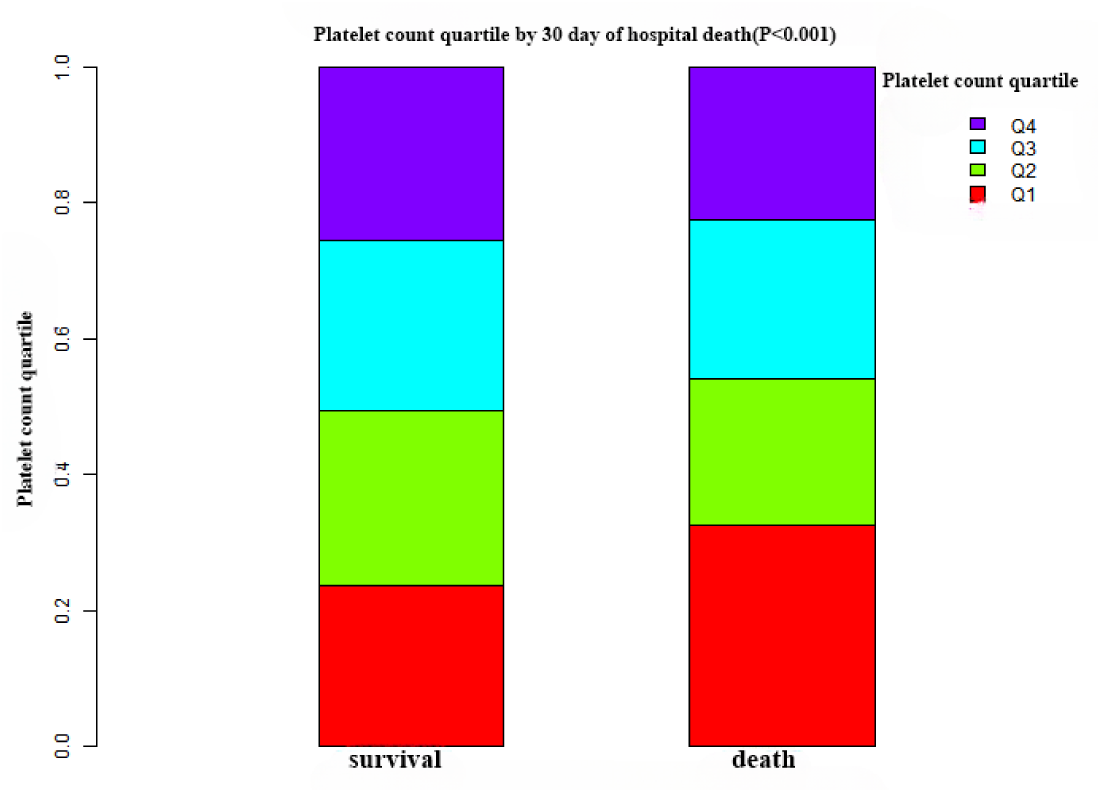
Platelet count levels in the survival and death groups. The figure illustrates lower platelet counts in the death group compared to the relatively higher platelet distribution in the survival group.

### 3.2. 30-Day In-Hospital Mortality Results

Table 2 presents the 30-day in-hospital mortality, indicating a rate of 14.02% (1151/8209). Notably, participants in the lowest quartile Q1 exhibited the highest mortality rate at 18.37% (95% CI: 16.69-20.05%), significantly surpassing the mortality rate in the highest quartile Q4, which stood at 12.51% (95% CI: 11.08-13.93%). Q3 displayed an intermediate rate of 13.26% (95% CI: 11.79-14.76%), aligning between Q1 and Q4. A discernible downward trend in mortality rates is observed from lower to higher platelet count quartiles, as supported by the significant P for trend 0.0001 (see Figure 4).

**Fig. 4:**
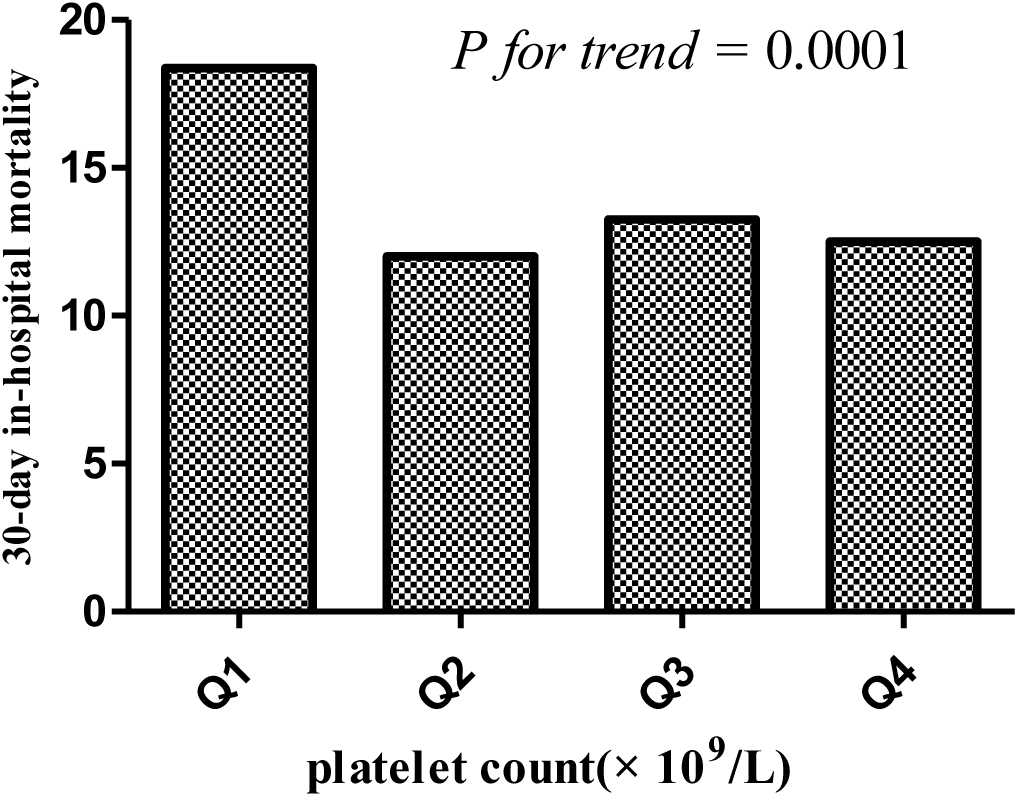
30-Day in-hospital mortality across platelet count quartiles.

### 3.3. Univariate Analyses Results Using a Binary Logistic Regression Model

Table S2 displays the outcomes of univariate regression analyses investigating the impact of various factors on 30-day in-hospital mortality. Gender showed no significant association with mortality, with females having similar odds (OR 1.01, 95% CI 0.89-1.14, P=0.8774) compared to males. Similarly, there was no substantial correlation between ethnicity and mortality (P>0.05). Elevated age (OR 1.01, 95% CI 1.01-1.02, P<0.0001) and APACHE-IV score (OR 1.04, 95% CI 1.04-1.04, P<0.0001) significantly predicted higher mortality. Conversely, a reduction in Hb levels (OR 0.94, 95% CI 0.91-0.96, P<0.0001) and a decrease in BMI (OR 0.99, 95% CI 0.98-1.00, P=0.0366) were associated with significantly higher mortality rates. Several comorbidities were linked to significantly higher mortality, including ARF (OR 4.51, 95% CI 3.94-5.16, P <0.0001), AF (OR 1.37, 95% CI 1.15-1.62), ACS (OR 1.87, 95% CI 1.45-2.40, p<0.0001), CHF (OR 1.83, 95% CI 1.44-2.31, P<0.0001), COPD (OR 1.52, 95% CI 1.17-1.99, P =0.0019), diabetes mellitus (OR 1.41, 95% CI 1.19-1.66, P <0.0001), and sepsis (OR 2.63, 95% CI 2.14-3.22, P <0.0001). The use of glucocorticoids (OR 1.82, 95% CI 1.39-2.37, P<0.0001), carbapenems (OR 2.11, 95% CI 1.12-3.97, P=0.0207), cephalosporins (OR 1.38, 95% 1.04-1.83), vancomycin (OR 2.56, 95% CI 2.00-3.29, P <0.0001), and mechanical ventilation (OR 7.27, 95% CI 6.35-8.31, P<0.0001) were associated with a higher mortality risk.

### 3.4. Multivariate Analyses Results Using the Binary Logistic Regression Model

Table 3 delves into the association between platelet count and 30-day in-hospital mortality through crude, adjusted, and fully adjusted logistic regression models. In the crude model, a 10-unit increase in platelet count correlated with a 2.9% lower risk of mortality (OR 0.971, 95% CI 0.963-0.979, P<0.0001). In the minimally adjusted model (Model I), where adjustments were made solely for demographic variables (age, sex, and ethnicity), each 10-unit rise in platelet count resulted in a 2.8% reduction in mortality risk (OR 0.972, 95% CI 0.964-0.981, P<0.0001). In the fully adjusted model (Model II), a 10-unit increase in platelet count was linked to a 2.5% decrease in mortality (OR = 0.975, 95% CI: 0.966– 0.984, P<0.0001). Significantly, in all three models, the mortality risk of Q2, Q3, and Q4 was lower than that in Q1, and the dose-response relationship remained significant (all P for trend<0.05). Additionally, we computed an E-value to evaluate sensitivity to unmeasured confounding, which was determined to be 1.21. This value, exceeding the relative risk of unmeasured confounders and platelet count, indicates that unknown or unmeasured confounders had minimal impact on the relationship between platelet count and 30-day in-hospital mortality.

### 3.5. Subgroup Analysis Results

We designated sex, ethnicity, ARF, AF, ACS, CHF, CKD, COPD, diabetes mellitus, GB, hypertension, sepsis, and mechanical ventilation as predetermined effect modifiers. Our objective was to observe the stability of the association between platelet count and 30-day in-hospital mortality across various subgroups. Table 4 displays the outcomes of interaction analyses and subgroup analyses exploring the connection between platelet count and 30-day in-hospital mortality. None of the interaction P values for the tested patient characteristics reached statistical significance, indicating the consistent treatment effect of platelet count on mortality across all subgroups.

### 3.6. Addressing Nonlinearity with Generalized Additive Models (GAM)

In this study, we employed GAM to explore potential nonlinearities in the relationship between platelet count and 30-day in-hospital mortality (refer to Figure 5). Following adjustments for various factors such as sex, ethnicity, age, BMI, Hb, Scr, APACHE-Ⅳ score, ARF, AF, ACS, CHF, CKD, COPD, diabetes mellitus, GB, hypertension, sepsis, anti-platelet, anticoagulant, glucocorticoid, carbapenems, cephalosporins, vancomycin, and mechanical ventilation, a significant nonlinear association emerged (Log-likelihood ratio test P <0.0001). Table 5 outlines the outcomes of a two-piecewise linear regression model examining the link between platelet count and 30-day in-hospital mortality. The model identified an inflection point at 163×10^9^/L platelet count. Below this threshold, each 10-unit increase in platelet count correlated with an 8% reduction in mortality risk (OR 0.92, 95% CI 0.89-0.95, P<0.0001). However, beyond 163×10^9^/L, the mortality risk plateaued, showing no significant change (OR 0.99, 95% CI 0.98-1.00, P=0.1169). The log-likelihood ratio test comparing the two models was highly significant (P<0.001), indicating that the two-piecewise model better captured the data.

**Figure 5.**
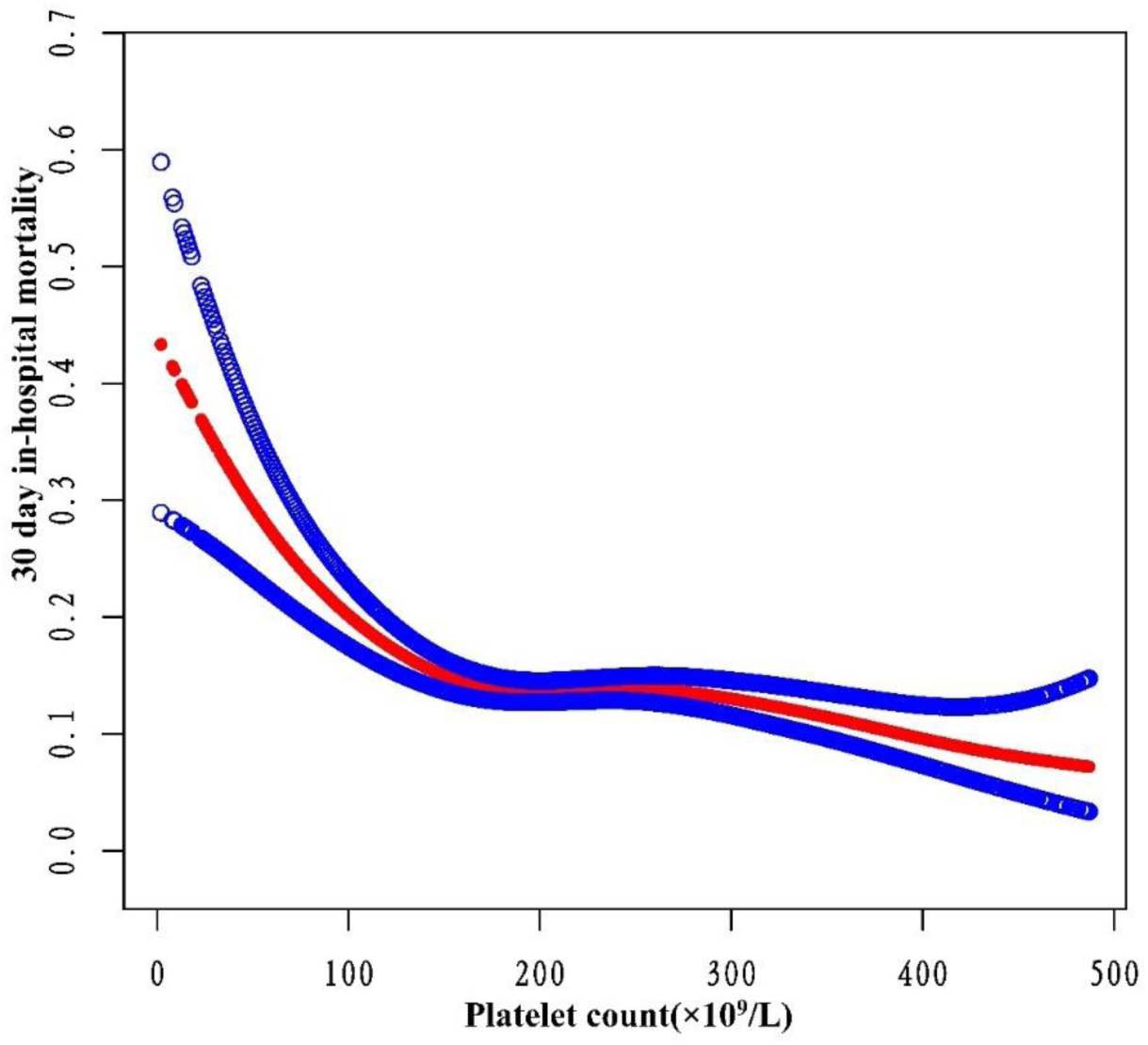
The nonlinear relationship between platelet count and 30-day in-hospital mortality. A nonlinear relationship was detected after adjusting for sex, ethnicity, age, BMI, Hb, Scr, APACHE-Ⅳ score, ARF, AF, ACS, CHF, CKD, COPD, Diabetes mellitus, GB, hypertension, sepsis, anti-platelet, anticoagulant, glucocorticoid, carbapenems, cephalosporins, vancomycin, and mechanical ventilation (Log-likelihood ratio test *P*<0.001).

## 4. DISCUSSION

This study utilized multicenter eICU-CRDv2.0 data to explore the association between platelet count and 30-day in-hospital mortality among ICU stroke patients. The findings indicate that higher platelet count are linked to a significantly reduced risk of mortality. Furthermore, this relationship remains consistent across various subgroups. Notably, a threshold effect curve reveals distinct correlations between platelet count and 30-day in-hospital mortality on either side of the inflection point. Below 163×10^9^/L, each 10-unit rise in platelet count is associated with an 8% reduction in 30-day in-hospital mortality. Beyond 163×10^9^/L, the increase in platelet levels ceases to significantly impact mortality.

The association between platelet count and adverse outcomes in critically ill patients has garnered increased attention. Jonsson AB et al., in a cohort study involving 215,098 ICU patients, discovered that thrombocytopenia was linked to elevated mortality (28). AnthonCT et al., in a prospective cohort study spanning 52 ICUs across 10 countries with 1,166 patients, observed that 43% of critically ill patients developed thrombocytopenia, leading to worse outcomes, including increased mortality (29). In a retrospective study involving 167 COVID-19 ICU patients, Zhu Y et al. revealed that thrombocytopenia not only correlated with respiratory function deterioration but also increased mortality in severe COVID-19 cases(30). These studies underscore platelet count as a crucial prognostic indicator for critically ill ICU patients. Notably, Lillemäe K et al.’s multicenter retrospective study, encompassing 4,419 patients with traumatic brain injury, established that early thrombocytopenia was associated with an elevated risk of death in ICU-treated traumatic brain injury patients(31). This further emphasizes the impact of thrombocytopenia on the prognosis of neurological diseases. Despite this wealth of information, there is a noticeable gap in literature regarding the relationship between baseline platelet count and the 30-day risk of in-hospital mortality in ICU stroke patients. Consequently, prompted by these observations, we sought to explore the correlation between baseline platelet levels and the 30-day in-hospital mortality risk in ICU stroke patients.

This study not only established an association between low baseline platelet count and increased mortality in ICU stroke patients but also identified a saturation threshold effect. Initially, the authors examined the linear relationship between baseline platelet count reduction and the 30-day in-hospital death risk in ICU stroke patients through univariate and multivariate logistic regression analysis. A higher platelet count demonstrated a significant and independent correlation with lower 30-day mortality in a dose-dependent manner (P for trend 0.0001). Additionally, the stability of this relationship was confirmed through subgroup risk analysis (all P for interaction >0.05). Furthermore, employing GAM and smooth curve fitting methods, the authors delved into the nonlinear relationship between platelet count and 30-day in-hospital mortality in ICU stroke patients. Clinically, this implies that among patients with severe thrombocytopenia, even modest increases in platelet count confer meaningful survival benefits. However, once the count surpasses approximately 163×10^9^/L, additional increments cease to impact prognosis. Recognizing this nonlinear relationship holds significant implications for optimizing transfusion thresholds across various clinical scenarios. In summary, the two-piecewise regression model revealed a clinically informative inflection point in the platelet-mortality association. Drawing on previous research, we posit that thrombocytopenia and the heightened 30-day in-hospital mortality share several underlying mechanisms. Firstly, platelets play a pivotal role in hemostasis and coagulation(11). A decrease in platelet count inherently escalates the risk of primary or secondary hemorrhage in vital organs, diminishing overall survival rates. In ICU settings, where severe organ hemorrhage can be fatal, this risk is particularly pronounced. Secondly, reduced platelet count serves as an indirect marker of increased illness severity(32–36). Patients with lower platelet count often necessitate intensified medical interventions, such as heightened use of vasoactive drugs, renal replacement therapy, and mechanical ventilation. As corroborated by Table 1 in this study, the Q1 group exhibited elevated creatinine levels, higher APACHE-Ⅳ scores, and a greater incidence of complications like gastrointestinal bleeding, sepsis, and acute respiratory failure-all indicative of a heightened risk of mortality (all P <0.05). Such patients are prone to extended ICU stays and elevated mortality risks. Furthermore, platelets are acknowledged as the first responders in innate immunity, engaging with pathogens, including bacteria and viruses, via various surface receptors(37–39). Operating as immune cells, platelets also interact with other immune entities like neutrophils, monocytes, dendritic cells, and lymphocytes(40). These factors suggest that patients with lower baseline platelet count may experience compromised immunity, elevating their risk of mortality. These factors likely contribute to the observed negative correlation between platelet count and 30-day in-hospital mortality on the left side of this study’s inflection point. Conversely, on the right side of the inflection point, this relationship loses statistical significance, possibly owing to worsening patient conditions and heightened influence of other confounding factors. Therefore, the authors assert that maintaining an appropriate platelet count is pivotal.

Our study boasts several strengths that significantly contribute to the depth and reliability of our findings. Notably, its multicenter and relatively large sample size enhance the generalizability of our results. The novelty of our research lies in its focus on ICU stroke patients, a unique population that has not been extensively studied in this context before. Additionally, our exploration of non-linearity adds a nuanced layer to our understanding of the relationship between platelet count and 30-day in-hospital mortality, surpassing the scope of previous studies. The use of multiple imputations to handle missing data ensures the mitigation of potential bias, maximizing statistical power. We further bolster the robustness of our findings through meticulous subgroup analysis and the calculation of E values. Beyond the academic realm, the practical implications of our study are heightened by the cost-effectiveness and accessibility of platelet count as a general laboratory measure, underscoring its applicability in diverse clinical scenarios.

This work, however, has certain limitations. Firstly, our study focused exclusively on Americans, necessitating further validation across diverse geographic and ethnic groups to generalize the findings. Secondly, the observational nature of our study design precludes establishing definitive causality. Despite this limitation, we meticulously adjusted for confounding factors, ensuring robustness through subgroup analysis. The calculated E-value also indicates that unmeasured confounders are unlikely to significantly impact the results. Thirdly, the absence of stroke classification data in our database hinders a detailed exploration of the relationship between platelet count and specific stroke subtypes. Notably, our study exclusively included ICU-hospitalized stroke patients, warranting verification for those with milder strokes. These considerations underscore the need for incorporating more detailed variables in the design of future studies.

In summary, this study reveals a negative and nonlinear correlation between platelet count and 30-day in-hospital mortality among ICU stroke patients. Notably, a threshold effect is observed, indicating a significant negative association when the platelet count is ≤163×10^9^/L. Conversely, when the platelet count exceeds 163×10^9^/L, this relationship becomes statistically insignificant. These findings offer valuable insights for clinicians, suggesting that maintaining the platelet count around 163×10^9^/L could potentially reduce 30-day in-hospital mortality in ICU stroke patients.

## Data Availability

The eICU Collaborative Research Database, a freely available multi-center database for critical care research. The data is accessible to the public after registration, with the collection process meeting the standards of the MIT Ethics Committee (No: 0403000206) and aligning with the 1964 Declaration of Helsinki.

## Author’s Contribution

Lan-xiang Wang: data analysis and manuscript draft; Ren-li Liu and Pan Zhou: critical revision of the manuscript for important intellectual content; Hao-fei Hu: study concept and design; Zhe Deng: administrative, technical, and material support and study supervision

